# Forecasting the scale of the COVID-19 epidemic in Kenya

**DOI:** 10.1101/2020.04.09.20059865

**Authors:** Samuel P. C. Brand, Rabia Aziza, Ivy K. Kombe, Charles N. Agoti, Joe Hilton, Kat S. Rock, Andrea Parisi, D. James Nokes, Matt J. Keeling, Edwine W. Barasa

## Abstract

**Background:** The first COVID-19 case in Kenya was confirmed on March 13^th^, 2020. Here, we provide forecasts for the potential incidence rate, and magnitude, of a COVID-19 epidemic in Kenya based on the observed growth rate and age distribution of confirmed COVID-19 cases observed in China, whilst accounting for the demographic and geographic dissimilarities between China and Kenya.

**Methods:** We developed a modelling framework to simulate SARS-CoV-2 transmission in Kenya, KenyaCoV. KenyaCoV was used to simulate SARS-CoV-2 transmission both within, and between, different Kenyan regions and age groups. KenyaCoV was parameterized using a combination of human mobility data between the defined regions, the recent 2019 Kenyan census, and estimates of age group social interaction rates specific to Kenya. Key epidemiological characteristics such as the basic reproductive number and the age-specific rate of developing COVID-19 symptoms after infection with SARS-CoV-2, were adapted for the Kenyan setting from a combination of published estimates and analysis of the age distribution of cases observed in the Chinese outbreak.

**Results:** We find that if person-to-person transmission becomes established within Kenya, identifying the role of subclinical, and therefore largely undetected, infected individuals is critical to predicting and containing a very significant epidemic. Depending on the transmission scenario our reproductive number estimates for Kenya range from 1.78 (95% CI 1.44 −2.14) to 3.46 (95% CI 2.81-4.17). In scenarios where asymptomatic infected individuals are transmitting significantly, we expect a rapidly growing epidemic which cannot be contained only by case isolation. In these scenarios, there is potential for a very high percentage of the population becoming infected (median estimates: >80% over six months), and a significant epidemic of symptomatic COVID-19 cases. Exceptional social distancing measures can slow transmission, flattening the epidemic curve, but the risk of epidemic rebound after lifting restrictions is predicted to be high.

## Introduction

First identified in Hubei province, China, capital Wuhan, there has been since December 2019 an ongoing epidemic of atypical pneumonia (COVID-19) caused by the zoonotic novel coronavirus (SARS-CoV-2) (Li, Q et al. 2020). As of 8th April 2020 there have been over 1.4 million clinically and/or laboratory confirmed cases of COVID-19 with over 70,000 deaths <u>worldwide</u>. This is the first coronavirus pandemic.

A number of sub-Saharan African countries, including Kenya, were at moderate to high risk of novel coronavirus importation, measured by volume of air travel arriving from infected Chinese provinces (Gilbert et al. 2020). As of 9th March8th April 2020, no East African country has had more than 200 confirmed cases of COVID-19, and there are few confirmed instances of local transmission in the region (World Health Organization 2020). However, the likelihood of a significant outbreak remains high, with potentially severe consequences for fragile regional health systems (Makoni 2020). The potentially high negative impact of a novel coronavirus outbreak in Kenya provides a strong motivation for forecasting studies of COVID-19 epidemic magnitude ahead of a serious outbreak under a number of plausible scenarios. This modeling study provides a baseline for continuous updating as improved data become available, e.g. time course of COVID-19 cases, updated mobility estimates, and the proportion and infectivity of asymptomatic infections, and new intervention strategies are proposed or implemented. It also provides the basis for studies of health service capacity; see accompanying article (Barasa et al, 2020).

SARS-CoV-2 is the third novel betacoronavirus to pose a global health threat since 2002, the other two being: (i) severe acute respiratory syndrome coronavirus (SARS-CoV; in 2002), and (ii) Middle East respiratory syndrome coronavirus (MERS-CoV; in 2012). Both previous novel coronaviruses (nCoVs) were zoonotic, and clinically similar in their disease presentation, however they differed in their subclinical manifestation. There was virtually no evidence of asymptomatic infections of SARS-CoV, or of “hidden” person-to-person transmission caused by asymptomatic/subclinical or preclinical cases (Anderson et al. 2004). In contrast, MERS-CoV behaved more like other commonly circulating human CoVs, with a substantial proportion of asymptomatic or preclinical infections (Cauchemez et al. 2013).

### Age-dependent symptomatic rate

The role of asymptomatic infected individuals (in this study we abbreviate to “asymptomatics”) in the transmission of SARS-CoV-2 remains unclear. A significant majority of detections of SARS-CoV-2 infection have occurred after infected individuals have presented clinically with COVID-19. Clinical cases of COVID-19 have presented mostly among older age groups (CPERE 2020). This could be due to a stronger resistance among younger age groups to contracting SARS-CoV-2 (Zhang et al. 2020; Hilton & Keeling 2020), or due to younger people developing COVID-19 symptoms at a lower rate after contracting SARS-CoV-2, leading to under-ascertainment amongst those age groups. One epidemiological study of 391 confirmed cases and their 1,286 close contacts in Shenzen found that children were as likely to be infected as adults (Bi et al. 2020), despite presenting with symptoms less frequently. Another modelling study emphasizes the role of undetected subclinical transmission in explaining the observed epidemic dynamics in China (Li, R. et al. 2020). These findings informed a central assumption of our modelling: that all age-groups are equally susceptible to SARS-CoV-2 infection but that the symptomatic rate is different across age groups.

In a recent novel coronavirus transmission modelling study by Wu *et al* (Wu et al. 2020) a clinical SARS-CoV-2 case was defined essentially tautologically: a clinical case is one that has sufficiently severe symptoms that the individual is detected by the health authorities. However, this presents a difficulty in translating symptomatic rates derived from Chinese confirmed COVID-19 clinical case data into a prediction of incidence in the Kenyan setting. Mildly symptomatic cases, defined as non-pneumonia and mild pneumonia cases by China CDC (CPERE, 2020), that presented to the health system in China might not present to the health system in Kenya. In this study, we provide forecasts of the possible epidemic course of COVID-19 infections, focussing on symptomatic infection, following initial invasion of SARS-CoV-2 into Kenya. We recognize the limitations of using clinically detected cases from another setting (China) to infer a symptomatic rate in this setting (Kenya). This reflects the difficulty in ascertaining COVID-19 disease with an unclear available testing capacity and coverage. This limitation highlights the importance of the sensitivity analysis in this modelling study. We treated all sub-clinical undetected infected individuals in China as asymptomatic. However, the threshold for being a sub-clinical infected individual will be different in Kenya and might miss significantly infectious, or even diseased, individuals. This observation led us to consider a range of infectiousness scenarios for asymptomatic cases, each scenario being capable of explaining the age-distribution of cases observed in China.

## Results

### Reproductive number predictions for Kenya

We assumed that the fundamental epidemiological characteristics of SARS-CoV-2 infection in Kenya would be like those estimated in China. These characteristics include: the incubation period, the infectiousness period, age-dependent symptomatic rate, and the per-contact transmission probability. A key epidemiological value to estimate is the reproductive number; the mean number of secondary infections, both symptomatic and asymptomatic, per typical infected individual. Despite using the same epidemiological characteristics as China, the reproductive number for Kenya will not be the same as that estimated for the early stages of the pandemic in China for two main reasons:

1. The age-profile of the Kenyan population is significantly different to that of China; Kenya having a far younger populace. Therefore, the symptomatic rate of a “typical” infected individual will be different in Kenya compared to China.
2. The typical mixing patterns between different age groups differ between the two countries. Therefore, the transmission rate will differ even if the per-contact transmission probability is the same.

Our central assumption is that all age groups are equally susceptible to SARS-CoV-2 infection and that differing rates of becoming symptomatic are responsible for the age distribution of confirmed COVID-19 cases. The age-dependent symptomatic rate could not be identified independently from an assumption about the infectiousness of undetected asymptomatics relative to detected symptomatic cases. Therefore, we inferred age-dependent symptomatic rates, using the age distribution of confirmed COVID-19 symptomatic cases in China, for a range of values of the relative infectiousness of asymptomatics to symptomatic infected individuals. The range of relative infectiousness scenarios considered in this study was 0%, 10%, 25%, 50%, and 100%. Each of these scenarios is capable of explaining the age-distribution of cases observed in China, but with a different set of fitted symptomatic rates in each scenario (Figure 1).

**Figure 1:**
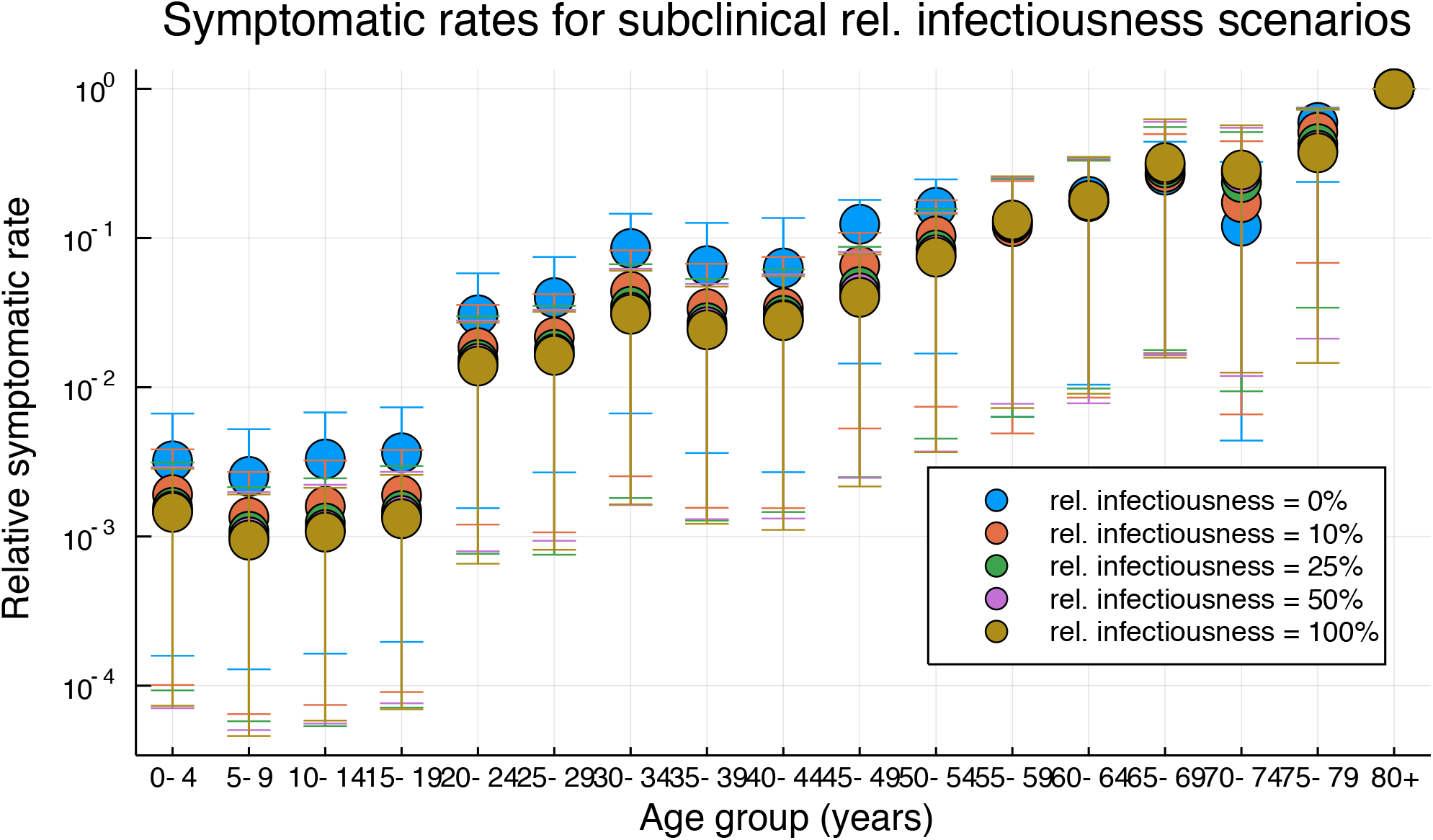
Estimates of symptomatic rates per age group relative to symptomatic rate for 80+ year olds.

It is also possible to explain the age distribution of cases observed in China by assuming a biological mechanism exists which reduces the susceptibility of contracting SARS-CoV-2 for some age groups compared to others. In the early stages of an epidemic, it is not possible to distinguish between age-differing susceptibility and age-differing symptomatic rate if asymptomatics are not infectious (0% relative infectiousness).

We find that our estimates of the Kenyan reproductive number (R_0_), without interventions, depend strongly on the relative infectiousness of the asymptomatic cases. As a baseline, if the symptomatic rate was age-independent, so that age-structured mixing was the only driver of age heterogeneity, then we would expect the reproductive number in Kenya to be 38.3% higher than in China, because of the typically higher mixing rates estimated within the Kenyan population compared to the Chinese population (Prem et al. 2017). In a scenario where younger age groups are developing COVID-19 symptoms at a lower rate after infection compared to older age groups and their infectiousness is negligible (0%) then the R_0_ we predict for Kenya is substantially lower than the R_0_ for China (28.8% lower). This is also true if younger age groups are more resistant to contracting SARS-CoV-2 (Table 1). However, in scenarios where asymptomatic infected are transmitting SARS-CoV-2 within the community the predicted R_0_ for Kenya is higher than the estimates for China, and this prediction holds even if the relative infectiousness of asymptomatic cases in Kenya is 10% that of symptomatic cases (Table 1).

**Table 1:**
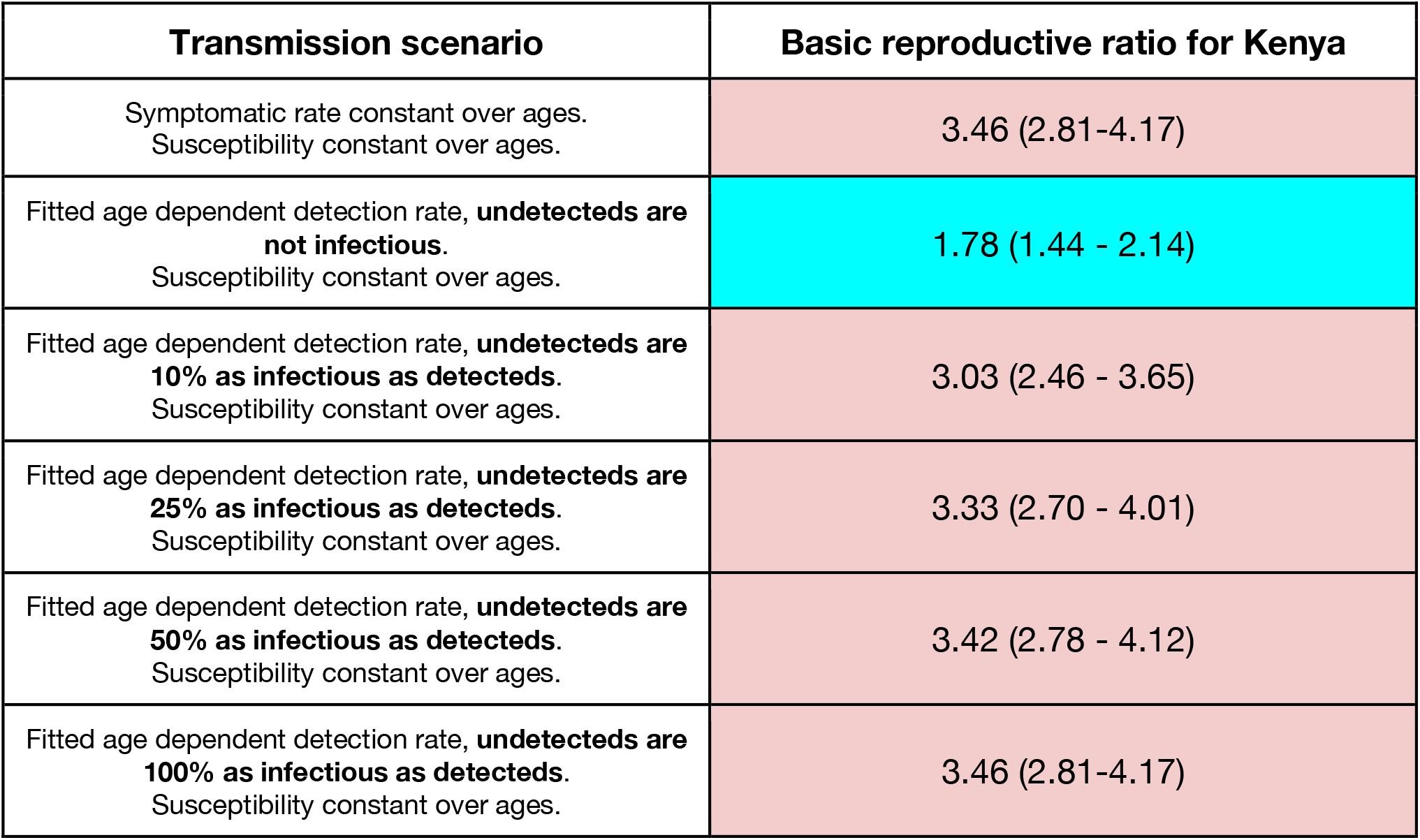
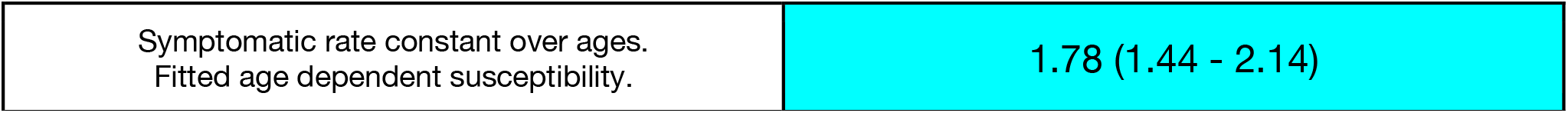
Reproductive ratio for Kenya for different transmission scenarios, before intervention. Based on a R_0_ for China of 2.5 (2.03 - 3.01). Color-coded for better (blue) or worse (red) than China R_0_.

| Transmission scenario | Basic reproductive ratio for Kenya |
| --- | --- |
| Symptomatic rate constant over ages.<br>Susceptibility constant over ages. | 3.46 (2.81-4.17) |
| Fitted age dependent detection rate, **undetecteds are not infectious**.<br>Susceptibility constant over ages. | 1.78 (1.44 - 2.14) |
| Fitted age dependent detection rate, **undetecteds are 10% as infectious as detecteds**.<br>Susceptibility constant over ages. | 3.03 (2.46 - 3.65) |
| Fitted age dependent detection rate, **undetecteds are 25% as infectious as detecteds**.<br>Susceptibility constant over ages. | 3.33 (2.70 - 4.01) |
| Fitted age dependent detection rate, **undetecteds are 50% as infectious as detecteds**.<br>Susceptibility constant over ages. | 3.42 (2.78 - 4.12) |
| Fitted age dependent detection rate, **undetecteds are 100% as infectious as detecteds**.<br>Susceptibility constant over ages. | 3.46 (2.81-4.17) |
| Symptomatic rate constant over ages.<br>Fitted age dependent susceptibility. | 1.78 (1.44 - 2.14) |

### Predictive modelling of SARS-CoV-2 transmission for Kenya

In this modelling study, we are investigating the course of a potential epidemic of COVID-19 cases in Kenya. We are focused on medium-to-long term predictions, assuming that detection at port-of-entry and within-Kenya contact tracing fails to suppress transmission immediately after introduction. The experience of countries which have experienced a substantial epidemic of COVID-19 cases has been that the onset of rapid growth in clinical cases, without any obvious transmission pathway between cases (Grasselli et al. 2020), was preceded by a period of largely undetected transmission in the community identified subsequently via genetic analysis of sequenced strains (Bedford et al. 2020). We recreate this “invisible” period of spread by assuming that there have been five generations of undetected transmission in Nairobi before the first set of Kenyan symptomatic cases are determined. In each of the scenarios presented below we present results starting from the exponential growth phase of the epidemic.

The underlying transmission scenario cannot be disentangled from the symptomatic rate estimate. We estimate that if the relative infectiousness rate of asymptomatics is higher, then their symptomatic rate must be lower to account for the observed case distribution in China (Figure 1). Consequently, the level of asymptomatic infectiousness in our model influences our expectation of the epidemic’s progress after five generations of unobserved transmission. In all scenarios, there were a small (1-6) number of initially symptomatic cases. In scenarios where the relative infectiousness of asymptomatics is higher, we are therefore also initializing with more asymptomatics to match the expected growth of the outbreak before detection.

In line with our reproductive ratio analysis, if asymptomatics were contributing a negligible amount of transmission (0% relative infectiousness) then in the absence of interventions we predict a comparatively long epidemic (median estimate being 8 months until new symptomatic incidence ceases, but with a high degree of uncertainty; Figure 2) with a slow growth rate of daily incidence, and the median estimate of daily symptomatic cases goes above 100 per day about three months after the detection of the first cases (Figure 2). Left unchecked we would expect an epidemic with 0% asymptomatic infectiousness to result in 1.06 million symptomatic cases of COVID-19 in Kenya among a 56.3% overall population infection rate, but with an exceptionally high level of prediction uncertainty (95% PI 0 – 1.44m symptomatic cases 0-75. 2% population symptomatic infection rate; Table 2).

**Table 2:** Predictions for final numbers of symptomatic cases and peak timing in two scenarios: unchecked epidemic and rapid isolation of first 1000 cases with 50% reduction in infectiousness of clinical cases throughout epidemic.

| Transmission scenario | Uncontrolled epidemic |  | Case isolation |  |
| --- | --- | --- | --- | --- |
|  | Total cases (millions symptomatic / % total infections) | Time of peak (days) | Total cases (millions symptomatic / % total infections) | Time of peak (days) |
| Fitted age dependent detection rate,<br>**Asymptomatics are not infectious.**<br>No age dependent susceptibility. | 1.06 (0.0, 1.44) | 109 (2, 217) | 0.0 (0.0, 0.0) | 5 (1, 33) |
|  | 56.3% (0%, 75.2%) |  | 0% (0%, 0%) |  |
| Fitted age dependent detection rate,<br>**Asymptomatics are 10% as infectious as detecteds.**<br>No age dependent susceptibility. | 1.02 (0.87, 1.13) | 65 (47, 93) | 0.94 (0.80, 1.08) | 68 (48, 97) |
|  | 88.4% (81.5%, 92.6%) |  | 85.8% (78.3%, 91.1%) |  |
| Fitted age dependent detection rate,<br>**Asymptomatics are 25% as infectious as detecteds.**<br>No age dependent susceptibility. | 0.915 (0.78, 1.02) | 58 (43, 85) | 0.88 (0.75, 0.99) | 60 (43, 88) |
|  | 89.0% (82.5%, 92.8%) |  | 87.9% (81.3%, 91.9%) |  |
| Fitted age dependent detection rate,<br>**Asymptomatics are 50% as infectious as detecteds.**<br>No age dependent susceptibility. | 0.89 (0.77, 0.99) | 56 (40, 81) | 0.87 (0.75, 0.98) | 57 (41, 83) |
|  | 89.1% (83.4%, 92.8%) |  | 88.5% (82.4%, 92.5%) |  |
| Fitted age dependent detection rate,<br>**Asymptomatics are 100% as infectious as detecteds.**<br>No age dependent susceptibility. | 0.87 (0.74, 0.97) | 56 (41, 82) | 0.86 (0.74, 0.97) | 56 (41, 80) |
|  | 89.0%<br>(82.7%, 92.7%) |  | 88.8%<br>(82.7%, 92.5%) |  |

**Figure 2:**
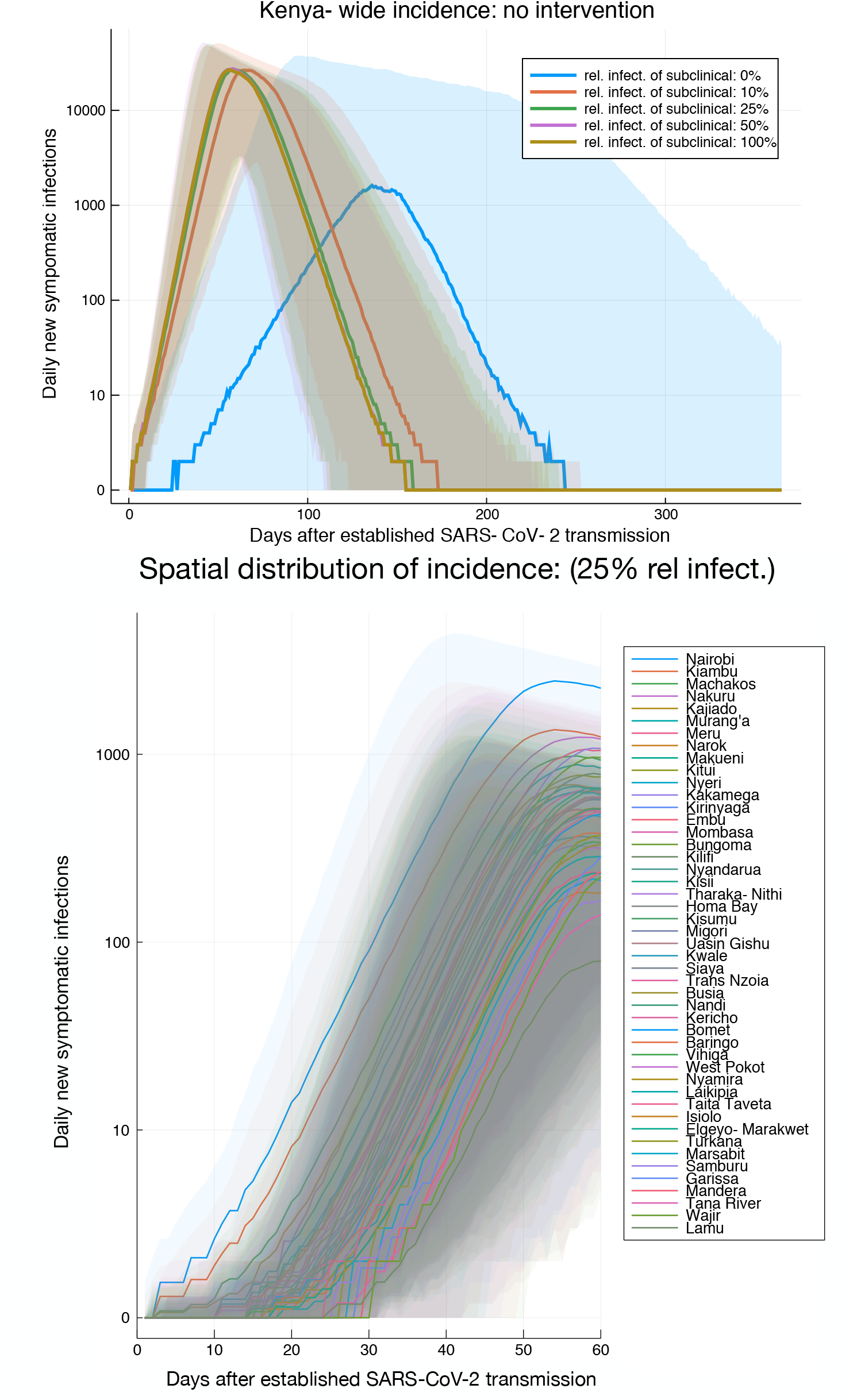
Baseline estimates of incidence rates and early spatial distribution of incidence. (*Top)* Daily incidence of symptomatic infections (median) estimates across Kenya for an uncontrolled epidemic and five different rates of subclinical transmission. Ribbon plot give median estimates with 95% prediction intervals. (*Bottom*) As *top* but incidence is given for each county using the 25% relative subclinical transmission scenario as representative. Legend is organized by median incidence rate at 40 days.

If asymptomatics are contributing to the epidemic, then we predict a substantially faster spread than in the 0% relative infectiousness scenario. Transmission amongst the younger age groups, who are expected to be largely asymptomatic, substantially boosts the early transmission rate within the Kenyan population, leading to a rapidly arriving peak (median estimates: < 3 months to peak, and 6 months until end of epidemic, after beginning of exponential growth in symptomatic infections) and a high probability of more than 10,000 daily symptomatic infections around the peak (Figure 2). In these scenarios, where asymptomatic infected individuals are contributing to transmission, and in the absence of effective intervention, we made a median estimate that novel coronavirus would spread first from Nairobi into the Lake Victoria regions and central regions. This spread is then closely followed by spread to Mombasa and throughout the coastal region. However, the uncertainty intervals on this prediction are substantial (Figure 2). Our median prediction is that, if unchecked, by 50 days after the establishment of SARS-CoV-2 circulating in the Kenyan population there will be more than 10 symptomatic cases of COVID-19 daily in every region of Kenya, but that this depends on essentially chance events early in the epidemic (Figure 2). For unchecked epidemics with asymptomatic transmission our estimates of total numbers of symptomatic infections over the course of the epidemic range from 1.02 million symptomatic infections, 88.4% population infection rate (95% PIs 0.87m – 1.13m, 81.5-92.6%) in the 10% relative infectiousness scenario to 0.86 million symptomatic infections, 89.0% total population infection rate (95% PIs 0.74m – 0.97m, 82.7-92.7%) in the 100% relative infectiousness scenario (Table 2).

Public compliance with government advice on self-isolation following the development of COVID-19 symptoms (50% reduction in the infectiousness of symptomatic cases) and rapid isolation of the first 1000 symptomatic cases (isolation achieved on average by 3.5 days post incubation) would be sufficient to contain a major outbreak in the scenario where undetected cases do not transmit (0% infectiousness), reducing the median predicted symptomatic infections from over 1 million to effectively zero if implemented immediately after first detection (Table 2). However, case isolation and reduction in infectiousness of the detected/clinical cases alone was insufficient to stop a major outbreak in any forecast simulation with asymptomatic transmission, or even achieve substantial delay in arrival of the peak or reduction in total cases (Table 2).

The essential difference in our predictions for the scenario with 0% relative infectiousness of asymptomatics and scenarios where asymptomatics are at least 10% as infectious as symptomatic infecteds could be seen in the different predicted age distributions of symptomatic cases. In the scenario with no transmission from asymptomatics the observed epidemic was dominated by cases among the working-age population (Figure 3), who we estimated as having high rates of assortative (i.e. within same age-group) mixing (Figure 4) and a small but not negligible risk of developing symptoms of COVID-19 after infection. In scenarios with at least 10% relative infectiousness of asymptomatics then the high rates of disassortative mixing between the elderly (75+ year olds) and children (Figure 4), who we expected to have a high rate of asymptomatic infection, led to the majority of observed cases occurring in the elderly population despite their relatively small proportion of the Kenyan population (Figure 3).

**Figure 3:**
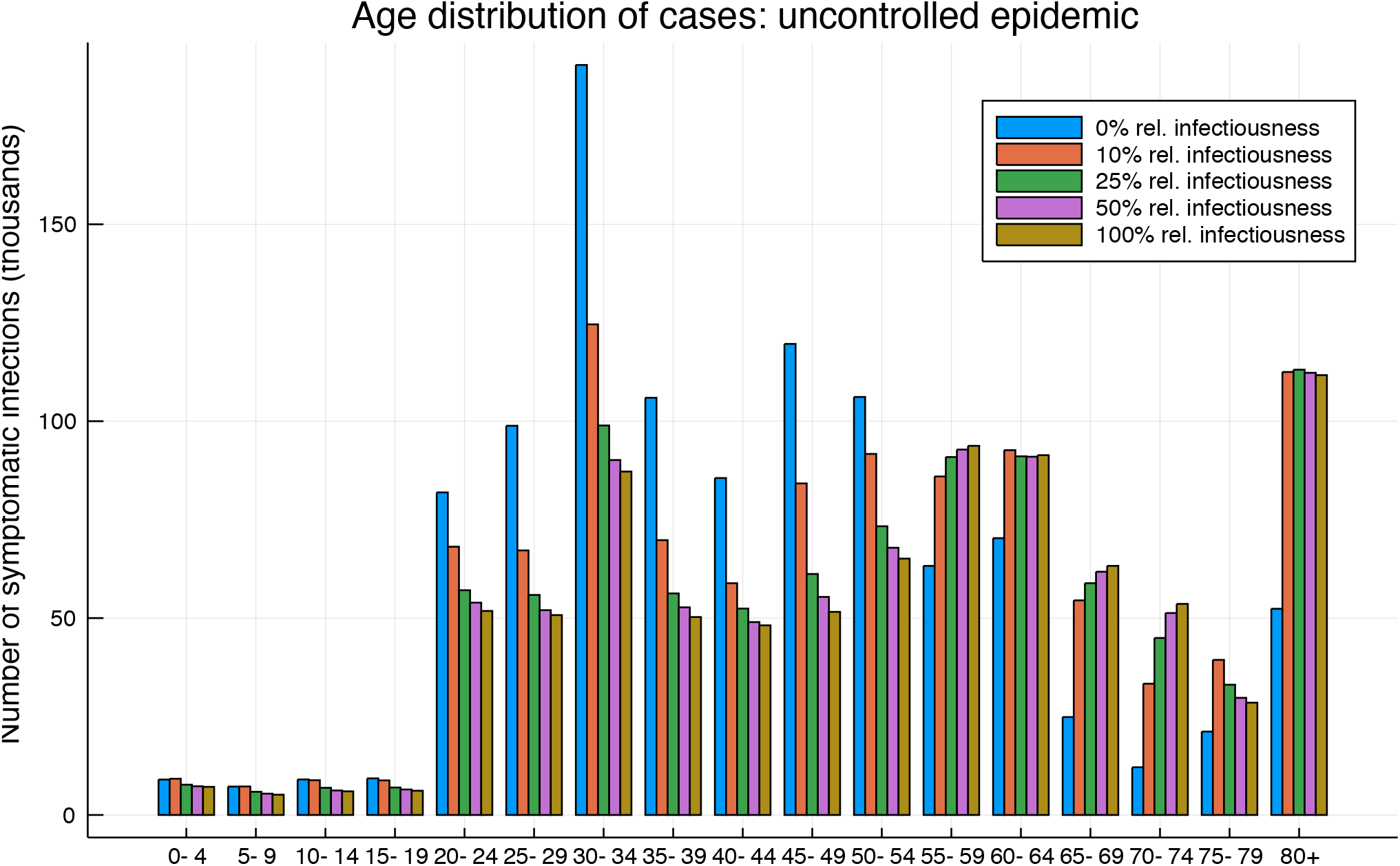
Distribution of symptomatic cases by age for all transmission scenarios. Median estimates of number of clinical cases by age group over the course of an uncontrolled epidemic.

**Figure 4:**
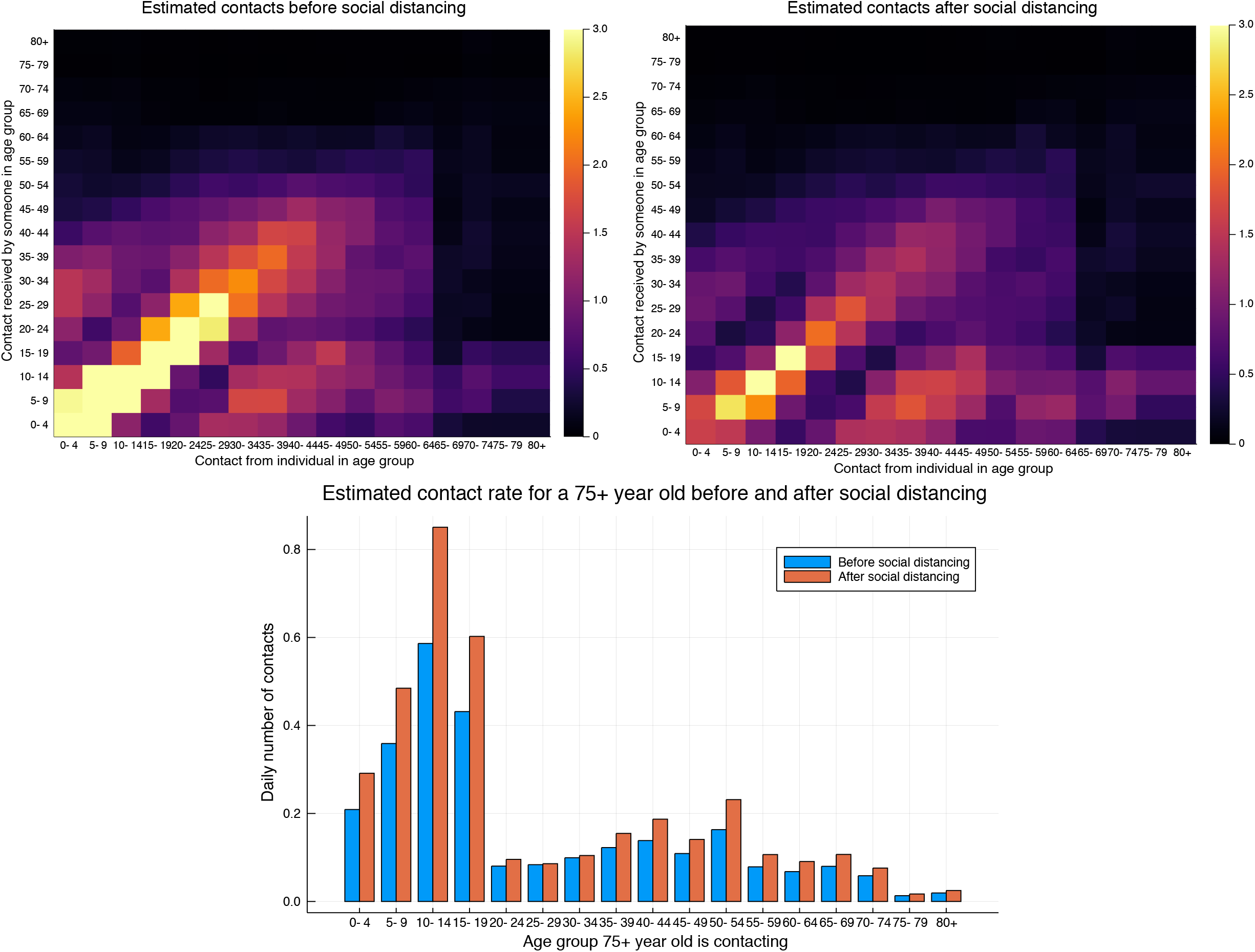
The predicted effect of social distancing on contact rates. (*Top left*) Daily rate of contact between an individual in an age group (column) and any other individual in some age group (row) before social distancing restrictions. (*Top right*) Daily rate of contact between an individual in an age group (column) and any other individual in some age group (row) immediately after social distancing restrictions. (*Bottom*) The rate of contacting other individuals by age group for a 75+ year old before, and after, social distancing measures.

In scenarios where asymptomatics are transmitting within the population without being detected, and person-to-person transmission has become established in Kenya, it is unlikely that case isolation alone will be sufficient to contain a Kenyan outbreak. Therefore, we also estimated the impact of wide-ranging social distancing (SD) measures and movement restrictions (MR) on the epidemic progression, in conjunction with case isolation (CI). In reality, an implementation of these measures would include school closures, a ban on mass gatherings, a sharp reduction in non-essential social contacts, and a sharp decrease in the amount of time individuals spend away from home. The estimated effect of social distancing was to reduce assortative social mixing among both children and working-age adults (Figure 4), thereby reducing transmission overall. However, we expected social contacts to be increased within the household as individuals spend more time at home. This had the effect of increasing the contact rate for 75+ year old individuals compared to before SD measures were implemented, because there is believed to be frequent within-household mixing between the elderly and children/young adults in Kenya (Figure 4).

This combination of interventions (SD+MR+CI) was predicted to be sufficient to slow down transmission and flatten the epidemic curve of COVID-19 in the scenarios where asymptomatics transmit SARS-CoV-2; that is, to eventually push the Kenyan reproductive number below 1. However, maintaining SD measures for the entire course of the epidemic is extremely challenging socially and economically. Therefore, we investigated the effect of implementing the full set of interventions for 90 days after the detection of established transmission within the country followed by a relaxation. We found that despite reducing transmission over the 90 days of implementation (median estimate 0 symptomatic cases after 90 days in all scenarios), there was a high probability of a rebound epidemic rapidly after relaxation of intervention measures. In the absence of a return to strict interventions if incidence rates are observed to increase, or novel effective vaccines/antivirals becoming available, the rebound epidemic was predicted to be similar in magnitude to the avoided epidemic (Figure 5).

**Figure 5:**
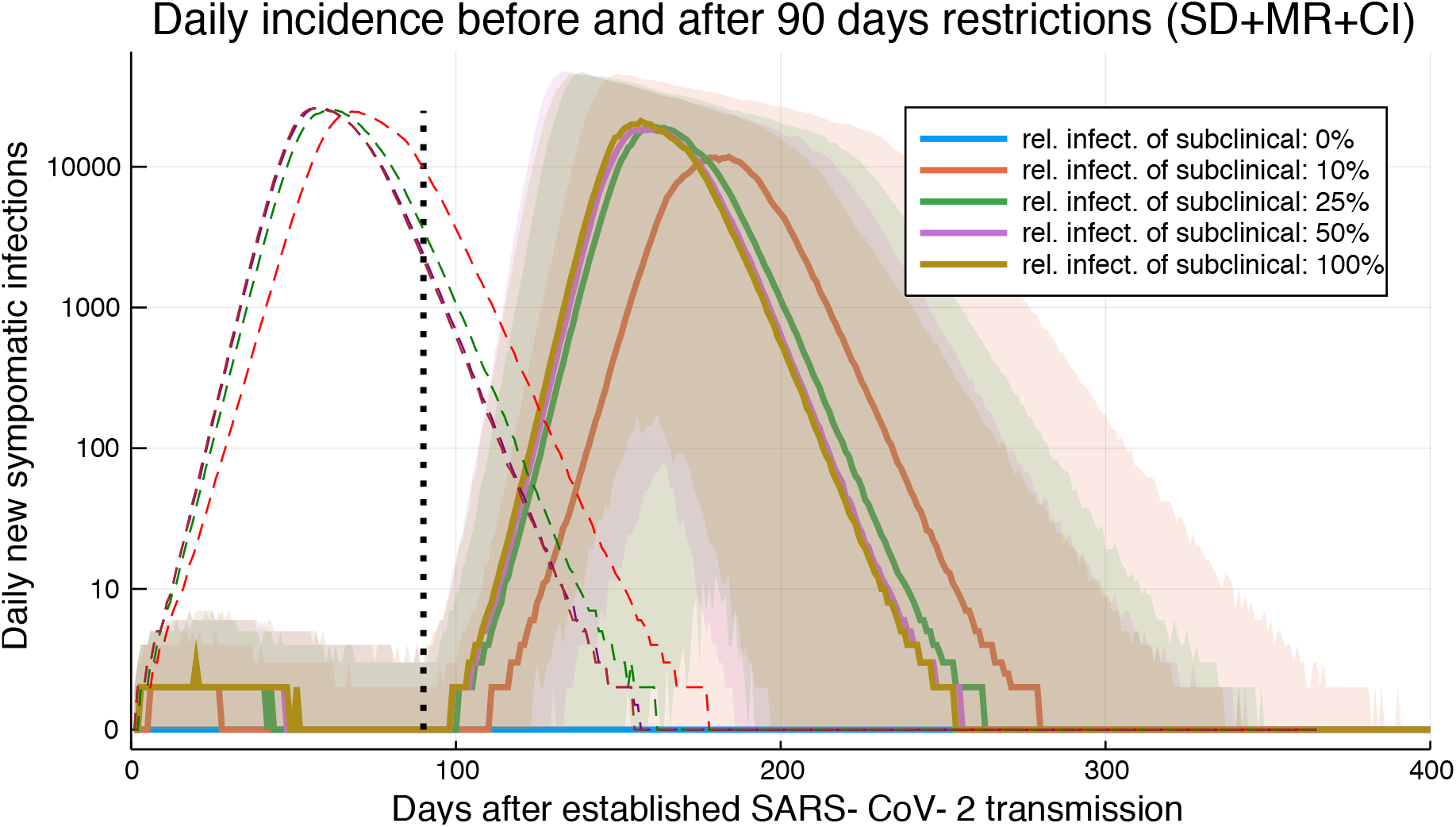
90 days of social distancing, movement restrictions and case isolation of first 1000 clinical cases. The daily incidence across Kenya by transmission scenario (as above ribbon plots give median and 95% PI). Full restrictions are assumed to apply from time zero until 90 days (black dotted line). The median predictions of daily incidence with only case isolation (CI), but no other intervention, are shown as dashed lines.

**Figure 6:**
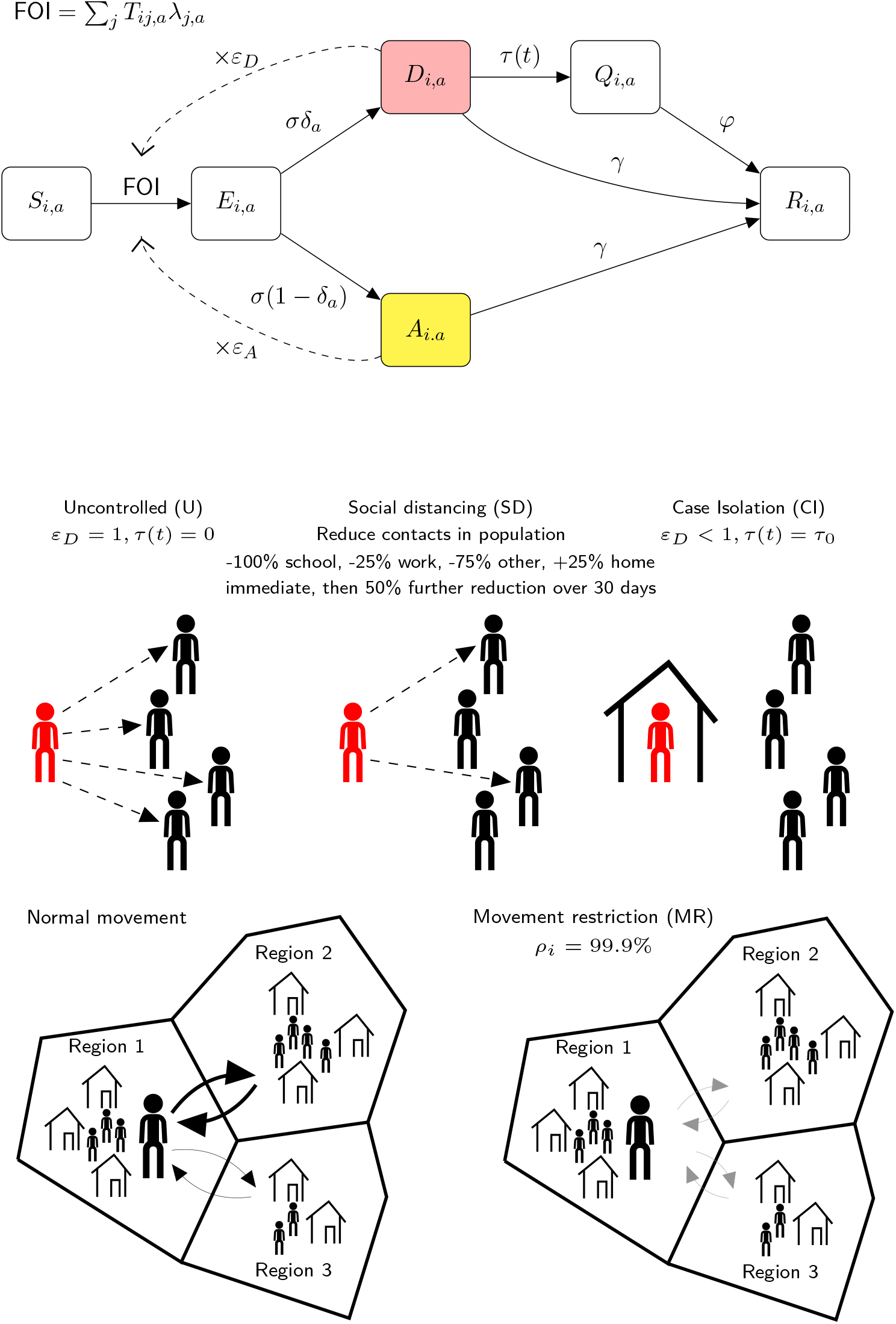
Schematic representation of KenyaCoV transmission and intervention modelling.

## Discussion

In this modelling study we have integrated existing data on the social structure, and mobility, of the Kenyan population with rapidly evolving estimates of the fundamental epidemiology of SARS-CoV-2 so as to make the best possible prediction of the scale of the epidemic risk that Kenya faces from the first coronavirus pandemic. We predict that the impact of an epidemic of COVID-19 cases in Kenya could be severe, with a high probability of observing more than 750 thousand infections with sufficient symptoms that they would have become clinical cases in the Chinese setting, if the epidemic was left unchecked. We predict that in the more pessimistic scenarios, and in the absence of an effective vaccine and/or anti-viral treatments, substantial restrictions on the social mixing and movement of the Kenyan people, either lasting for longer than three months, or reimplemented if incidence rebounds, would be required to control a COVID-19 epidemic in Kenya.

In this study we do not attempt to estimate the proportion requiring hospitalization, i.e. those having most impact on the health services, and most at risk of dying – nor estimate deaths. Estimating the true impact of the COVID-19 epidemic in Kenya requires an estimate of the clinical fraction of infecteds, the likelihood of clinical cases being severely ill and a detailed, and spatially explicit, understanding of the capacity of the Kenyan health service. Early work to assess Kenya’s health system capacity to absorb likely COVID-19 cases, using the predictions from this model on the likely scale of the pandemic, reveal that Kenya’s health system is likely to be overwhelmed because of limited availability oxygen and other essential devices to facilitate oxygen therapy (such as pulse oximeters), and critical shortages of intensive care beds (ICU) and ventilators (Barasa 2020). The analysis reports variation in Kenya’s surge capacity for hospital beds, with the county with the least surge capacity likely to need 145% of its available hospital beds to accommodate COVID-19 cases if the pandemic lasts for 6 months (Barasa 2020). The analysis shows that only 22 out of the 47 counties have at least one ICU unit and that the country will need an additional 1,511 ICU beds and 1,609 ventilators if the epidemic will last for 6 months. The authors indicate the need to strengthen lower cost essential services such as oxygen availability before focusing on these higher cost investments.

The extrapolation of the symptomatic rate from Chinese clinical observations to the Kenyan setting is likely to be imprecise, and should be replaced by symptomatic rates estimated from Kenyan case data as this becomes available. The Chinese epidemiological reports include a substantial proportion (80.9%) of mildly diseased cases (CPERE 2020); it is possible that a proportion of mildly diseased individuals in the Kenyan setting would not seek medical attention, and therefore would remain invisible to Kenyan surveillance. This observation suggests that the symptomatic rate estimated here could be an overestimate of the clinical incidence that will be seen in Kenya. On the other hand, it should be noted that our estimate for the symptomatic rate is somewhat lower in some age groups than other estimates in the literature. For example, Davies et al (Davies et al. 2020) make a consensus estimate of the clinical fraction of 0-9 year old infecteds as 20%, whereas we find the symptomatic rate for that age group is likely to be less than 1%.

As with all modelling studies, there are limitations in our modelling structure that go beyond uncertainty in parameterization. In particular, it is possible that controlling a COVID-19 epidemic in Kenya will require measures which can break transmission from household to household. However, an explicit household structure is not part of the KenyaCoV modelling framework. We replicate the effect of household structure in our modelling of social distancing using a combination of social context dependent age-structured mixing matrices and an effective decline in contact rate as individuals reduce contacts among the general population in favour of repeated contacts with a relatively small group of people within their own household. Whilst this is less realistic than a full, individual-based stochastic model, see for example Ferguson et al (Ferguson et al. 2005), the benefit of using KenyaCoV is that, in the event of a substantial COVID-19 epidemic in Kenya, the model is sufficiently performant that incorporating new information into forecasting could occur in near real-time. As mentioned above, it is critical that estimates of the symptomatic rate, clinical fraction, and rate of severe disease are derived from Kenyan case data rather than simply extrapolated from other settings. Such Kenyan-specific estimates of COVID-19, augmented by more recent data on Kenyan mobility, will allow us to better understand the link between transmission predictions (as given by KenyaCoV), and the observed clinical burden of COVID-19 in Kenya. Without understanding the link between transmission and clinical burden the design of public health interventions is done blind, and it is impossible to evaluate intervention success.

There remains a very high degree of uncertainty surrounding the underlying epidemiology of SARS-CoV-2 infection, which inevitably degrades our certainty in the predictive modelling of transmission in Kenya. We have attempted to partially account for these underlying uncertainties by drawing some parameters from a plausible range rather than simply using point estimates. However, some of the epidemiological uncertainties are fundamental to the structure of the KenyaCoV model. First, we treat the rate of developing symptoms as the detection rate in the model, following that the substantial majority of confirmed cases in the China were initially syndromic detections before test confirmation (CPERE 2020). However, since the symptomatic rates used in KenyaCoV were inferred from the age distribution of Chinese confirmed cases, this means that our model is predicting the incidence rate of cases *that would have been clinical in the Chinese epidemic*. It could be that the syndromic threshold for detection is different in Kenya, which would alter the effective transmission rate of the undetected cases by changing the definition of detected and undetected cases so that infectious and diseased individuals might be missed in the Kenyan context who would have been identified in the Chinese context. Second, and following from the previous comment, the rate of subclinical undetected transmission is critical to the expected outcome of an epidemic in Kenya. This reflects the fact that COVID-19 disease has been predominantly observed amongst older age groups, and therefore two major possibilities for a country with a younger demography arise. In the first possibility, the relatively large proportion of younger people effectively protect the elderly at-risk groups from transmission. In the second possibility, high rates of unobserved transmission within the young population cause significant numbers of clinical cases among the elderly. We don’t know which outcome will occur until a substantial number of cases are observed, and so far, no major COVID-19 epidemic has occurred in sub-Saharan Africa. If there exists climatic incompatibility for this strain of coronavirus and/or inherent resistance in the population then all modelling estimates derived from the Chinese experience should be revised downwards.

Finally, our model presents a partial picture of the likely effects of government interventions to slow the transmission of COVID-19. Government interventions such as social distancing and movement restrictions have secondary health, social and economic effects that should be considered when making decisions about how to intervene (Dahab et al. 2020; Barasa 2020). For instance, these interventions could disrupt routine health service delivery and access (such as vaccinations, and maternal and child health services), cause income and job losses in ways that could push households into poverty, and lead to a shrinking of country economies. Incorporating these dimensions in models, and ascertaining net health and non-health benefits of interventions would provide a fuller picture and improve the quality of decision making.

## Methods

### Data sources and assumptions

We parameterized our SARS-CoV-2 transmission model from a range of data sources. Epidemiological parameters for SARS-CoV-2 were collected from estimates that have been reported in the literature (Table 1). We followed Wesolowski *et al* (Wesolowski et al. 2012) in dividing Kenya into 20 regions corresponding to population density concentrations. These regions overlap the 47 counties of Kenya, and results for the 47 counties were created from simulations on 20 regions by apportioning infections pro-rata according to the proportion of the region population from each county.

We subdivided the population by their home region, and their age category. In this study we consider 17 age categories: 0-4 years old, 5-9 years old, …, 75-79 years old, 80+ years old. The modelled population size in each region/age category was chosen to match the recent <u>2019 Kenyan census</u> findings (denoted *N*_*i,a*_ for the number of individuals in region *i* and age group *a*) (KNBS, 2020).

We estimated the human mobility flux using mobile phone data on number and duration of trips from home regions to alternate regions (Wesolowski et al. 2012), rescaled to number of journeys per person per day. This allowed us to estimate both: (i) the proportion of time typical individuals in each region spent away from their home region (denoted *ρ*_*i*_ *∈* [0,1] for each region *i*), and (ii) a probability distribution for where typical individuals travel to when they leave their home region (denoted *P*_*i j*_ for probability of a person living in region *i* travelling to region *j*). The central mobility estimates depended on a reported median 5 nights duration per journey, which is likely to be an overestimate for short-distance journeys and an underestimate for long journeys. If the short-distance journeys are typically shorter than 5 nights, and the long-distance journeys are typically longer then the serial interval of SARS-CoV-2 then this central mobility estimate will overestimate the rate at which SARS-CoV-2 spreads spatially in Kenya.

The rate of contact between individuals currently in the same region, including visitors to the region, was determined by their age group using an estimated age-mixing contact matrix for Kenya from the study by Prem *et al* (Prem et al. 2017). Prem *et al* give estimated contact rates between age groups for 152 countries broken down by contact setting: at home, at work, at school, and other contacts amongst the population.

The epidemiological characteristics of SARS-CoV-2 were drawn from the literature and summarized into a belief distribution for key uncertain parameters (Table 3). The age-profile of cases used to infer the age-dependent symptomatic rates were from the first 44,672 confirmed cases as published by China CDC on 11^th^ February 2020 (CPERE 2020).

**Table 3:** Published and preprint estimates of SARS-CoV-2 epidemiological parameters used as evidence in this study.

| **Epidemiological parameter:**<br>**SARS-CoV-2** | **Estimates (95% CI)** | **Modelling belief distribution** |
| --- | --- | --- |
| Basic reproductive ratio ($R_0$) | $R_0 = 3.8$ (3.6-4.0) from early identified cases in Wuhan (Read et al. 2020).<br>$R_0 = 2.2$ (1.4-3.9) from the first 425 confirmed patients (Q. Li et al. 2020).<br>$R_0 = 2.68$ (2.47-2.86); from internationally exported cases until Jan 28th 2020 . $R_0 = 2.6$ (1.5-3.5); based on cases in Wuhan by 18th Jan 2020 (**Imperial group report**). | $R_0 \sim \text{Gamma} \left( \alpha = 100, \theta = \frac{2.5}{100} \right)$, |
| Mean incubation period ($1/\sigma$) | $1/\sigma = 5.2$ days (4.1-7.0) from first 425 confirmed patients (Q. Li et al. 2020).<br>$1/\sigma = 4.8$ days (no CI) (Liu et al. 2020). | $1/\sigma \sim \text{LogNormal}(\mu = \ln(5), \sigma = 0.25)$ |
| Mean serial interval | Mean serial interval = 7.5 days (5.3-19) from first 425 confirmed patients (Q. Li et al. 2020). | 7.5 days (point estimate) |
| Mean infectious duration ($1/\gamma$) | Estimated as (mean serial interval – mean incubation period) | 2.5 days (point estimate) |
| Age dependent symptomatic rate ($\delta_a$) | Relative symptomatic rate fitted to Chinese case data (CPERE 2020) | Used posterior mean estimators for relative symptomatic rates. True |
|  |  | symptomatic rate for 80+ year olds<br>assumed to be 90%. |

### Spatial- and age- structured stochastic transmission of novel coronavirus and intervention modelling

We used a variant of the susceptible-exposed-infectious-recovered (SEIR) metapopulation model (Keeling & Rohani 2008), dubbed **<u>KenyaCoV</u>**, to simulate transmission of SARS-CoV-2 in Kenya. The code and open source data for KenyaCoV can be found at https://github.com/SamuelBrand1/KenyaCoV.

Individuals divided their time between their home region and other regions according to their age and the region-specific travelling patterns described above. For simplicity, we assumed that age groups of individuals younger than 16 and older than 49 didn’t move around the country, therefore we defined the *Transport matrix, T*_*ij,a*_, as

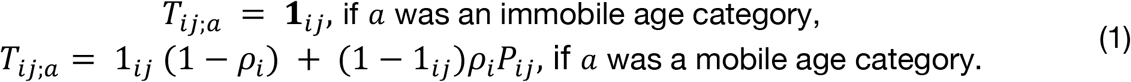

Where 1_*i j*_ *=* 1 if *i = j* and 0 otherwise. The transport matrix defined the proportion of time spent in each region.

The force of infection (the rate at which each susceptible becomes infected) on individuals in region *i* and age group *a* depended on their movements and age-dependent social contact rates. The force of infection on each susceptible individual **currently in region** *j* and age group *a* was,

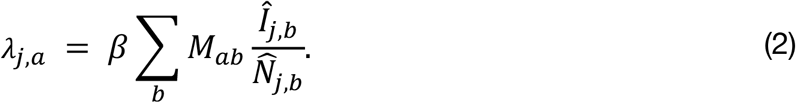

Where *β* was the SARS-CoV-2 transmission rate, *M* was the Kenyan age-mixing matrix, and *Î*_*j,b*_ and 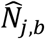 were, respectively, the *effective* number of infected individuals in, and the *effective* population size of, age group *b* currently in region *j*. Using effective numbers accounted for mobility of individuals and differential transmissibility between clinical cases and asymptomatics,

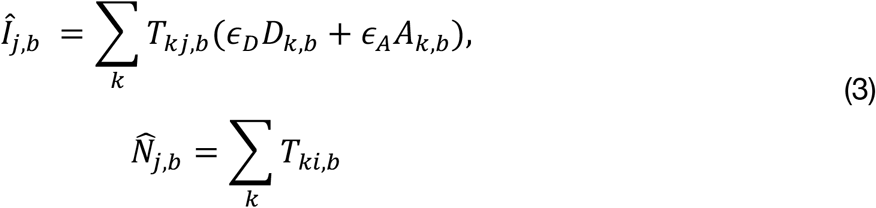

Where *ϵ*_*A*_ is the infectiousness of a asymptomatic infected relative to a clinical case, and *ϵ*_*D*_ was the reduction in infectiousness of clinical cases after they developed symptoms.

When individuals are infected with SARS-CoV-2, and have finished their incubation period, then they **either** develop sufficient symptoms to eventually become a clinical case (with age-dependent probability *δ*_*a*_) **or** they are sufficiently asymptomatic to remain subclinical and remains undetected (with probability 1 − *δ*_*a*_).

**The model treats population groups as discrete, and follows stochastic dynamics,** however, the overall KenyaCoV dynamical structure is most compactly represented using notation for an ordinary differential equation (ODE), the transition rates given in equation (4) should be interpreted as stochastic rates for each event,

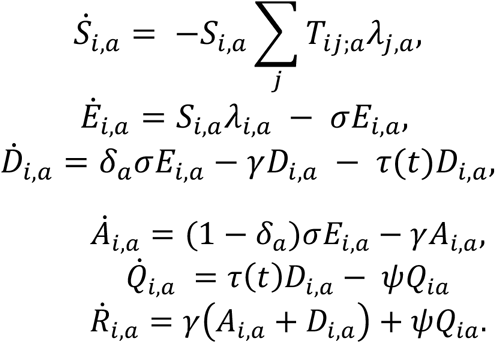

Where S_*i,a*_, *E*_*i,a*_, *D*_*i,a*_, *A*_*i,a*_, *Q*_*i,a*_, and *R*_*i,a*_ were the number of susceptible, latent, infectious with sufficient symptoms to become clinical cases, infectious but subclinical, isolated and removed individuals whose home was region *i* and age group. α, β, σ γ, Ψ were, respectively, the per-contact transmission probability, the incubation, recovery rate and exit rate from isolation. Note that the transmission structure included both the possibility that infection is spread around the country by the movements of infected individuals, and, by susceptible individuals who contract SARS-CoV-2 elsewhere and return to their home region.

### Inferring age-dependent symptomatic rates and the per-contact transmission probability

We inferred age-dependent symptomatic rates for each age group by matching the age distribution of clinical cases expected from combining the next-generation matrix for SARS-CoV-2 with symptomatic rates to the observed age distribution of clinical cases in China on 11^th^ February.

We denote the mixing rate at which individuals in age group *a* contact individuals in age group *b*, estimated for China, (*M*^*china*^)_*ab*_, as estimated by Prem et al (Prem et al. 2017). We use two versions of the age mixing matrix for China in this analysis: *M*^*china*^, which denotes the age mixing matrix including contacts in all social settings, and, *M*^*china,home*^, which denotes the age mixing matrix including contacts *only at home*.

For fitting age specific symptomatic rates we used *M*^*china,HO*^. Our reasoning was that the majority of cases reported in China occurred during severe lockdown restrictions on social contact outside of the household. The next-generation matrix across age groups (expected number of infections generated amongst age group *b* per infected in age group *a*) for China during lockdown was:

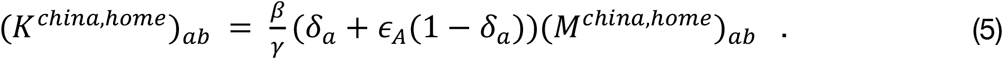

We denote the leading eigenvector of *(K*^*chinna,home*^)_*ab*_, as *C*_*a*_, normalized so that ∑_*a*_ *C*_*a*_ *=* 1. *C*_*a*_ is the expected proportion of infections amongst age group *a*, during the early stages of an outbreak but after a few generations of transmission (Diekmann & Heesterbeek 2000). The probability that amongst a collection of confirmed cases any given case is in age group *a* is *p*_*a*_ *= δ*_*a*_ *C*_*b*_/ ∑_*b*_ *δ*_*b*_ *C*_*b*_. This allowed us to infer a posterior distribution (we assumed flat priors) for the values of *δ*_*a*_ using Hamiltonian MCMC with a standard multinomial likelihood of recreating the observed case data in China. This was implementing using the **DynamicHMC** package for the Julia programming language. Note that *C*_*a*_ depends on *δ*_*a*_ and *ϵ*_*a*_, but not the per-contact transmission probability or the recovery rate. Similarly, *C*_*a*_ does not change is *δ* is scaled by a constant. Therefore, we fixed *δ*_17_ *=* 0.9, that is we assumed a true symptomatic rate amongst 80+ year olds of 90%.

For inferring the per-contact transmission probability, *β*, we matched to R_0_ estimates in China using the full Chinese contact matrix *M*^*china*^ by considering the leading eigenvalue of the **pre-lockdown** next generation matrix. This was chosen to reflect that we are using early estimates of R_0_ before lockdowns were fully achieved. Our estimated R_0_ for Kenya, without interventions, was the leading eigenvalue of *Kenyan* next-generation matrix using the Kenyan age-mixing matrix *M*_*ab*_.

### Modelling Interventions

We consider three types of interventions aimed at restricting SARS-CoV-2 transmission in Kenya:

- **Case isolation and reduction in transmission of clinical cases pre-isolation (CI)**. Clinical individuals are isolated from the rest of the population at an isolation/treatment rate *τ(t*), which represented the active intervention of the Kenyan health system and has an initial rate *τ(*0) *= τ*_0_. We assumed that if the cumulative number of detected cases reaches 1,000 individuals (∼0.002% of the Kenya population) then the health system has reached its capacity threshold and active isolation ceases (*τ(t*) *=* 0 after capacity is reached). Both before and after capacity is reached, the infectiousness of the clinical cases was reduced by a factor *ϵ*_*D*_ ≤ 1 which reflected passive intervention, e.g. public campaigns to raise awareness of SARS-CoV-2 risk, encourage self-isolation and social distancing of symptomatic individuals, and increased hygiene.
- **Wide-spread social distancing (SD)**. The whole Kenya population is required to minimize unnecessary social contacts, along with closing schools and colleges. We modelled this as decreasing work contacts by 25%, decreasing other contacts by 75%, offset by an increase in home contacts by 25%. We modelled the action of SD on the contact structure of the population by replacing the age-mixing matrix by a SD-mixing matrix,

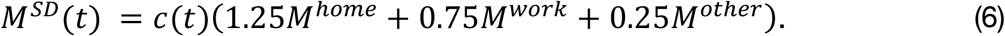 Where the three matrices *M*^*home*.^, *M*^*work*^, *M*^*other*^ are the estimated age-structured contact rates in, respectively, the home, work, and other settings (Prem et al. 2017). We followed Tang et al (Tang et al. 2020) in including a decreasing rate of contacts over time. The decreasing rate of contacts was combined with the change in age structured mixing, and attempted to account for increasing population compliance, a staggered step of extra interventions (e.g. closing non-essential shops and instigating a curfew), and the effective decrease in contacts across the population due to redirecting social contacts towards repeated contacts within the home. Specifically, we use the down-ramp function to represent a decrease in contacts by 50% over 30 days after the social distancing intervention: *c(t*) *=* 1 − 0.5*(t* – *t*_*I*_)/30 for times between the intervention time *t*_*I*_ and 30 days after SD intervention
- **Movement restrictions (MR)**. The whole Kenyan population is required to reduce movement around the country, with waivers only for specific reasons e.g. movement of vital supplies, movement of medical professionals. We model this by replacing the transport matrix with a post-movement restriction transport matrix, *T*^*MR*^, constructed by assuming that during movement restrictions 99.9% of the time of typical individuals was spent in their home region.

### Scenario forecasting for the spread of novel coronavirus in Kenya

We consider a range of possible levels of infectiousness of asymptomatics relative to individuals with severe symptoms who become clinical cases (*ϵ =* 0, 0.1, 0.25, 0.5, 1.). For each relative infectiousness scenario, detection rate, we ran 1000 simulations. We aimed to estimate the distribution of SARS-CoV-2 epidemic outcomes in Kenya over uncertainty in the fundamental epidemiological parameters. We achieved this by, prior to each simulation, drawing mean incubation period and reproductive ratio for China from a belief distribution of the parameter value that encompassed published literature estimates (Table 3). The Chinese *R*_0_ *∼ Gamma(α =* 100, *θ =* 2.5/100), which corresponded to a median belief that the SARS-CoV-2 *R*_0_ for China is equally likely to be above 2.5 as below (2.5-97.5 percentiles of belief *R*_0_ *=* 2.03 − 3.01). The mean incubation period 1/*σ ∼ LogNormal(μ = ln (*5), *σ =* 0.25); median belief 1/*σ =* 5 days (3.1-8.2 days). We fixed the mean infectious duration as 1/*γ* = 2.5 days, corresponding to the difference in the median serial interval less our median belief of the mean incubation period (Table 3).

A key point is that our belief distribution *R*_0_ is based on estimates derived from data of Chinese cases. Therefore, the transmission rate *β* used in each simulation, and which corresponds to some *R*_0_ drawn from our belief distribution, is derived using Chinese age-mixing data.

## Data Availability

All the data used in forecasts, and all simulation code, is available at KenyaCoV repository https://github.com/SamuelBrand1/KenyaCoV

## Acknowledgement

This paper was published with the permission of the Director of KEMRI.

